# Biological, experimental and analytical determinants influencing bile acids concentrations in human blood: a review and meta-analysis

**DOI:** 10.1101/2025.01.12.25320430

**Authors:** Sebastian Joseph, Sophie de Buyl, Isabelle A. Leclercq, Laure-Alix Clerbaux

## Abstract

**Background:** Despite over three decades of research, the use of peripheral bile acid concentrations or proportions as biomarkers for human liver injury remain inconclusive due to variable and inconsistent findings.

**Objective:** The aim of this systematic review and meta-analysis was to identify factors contributing to the variability in published bile acid research and propose recommendations to enhance the robustness and reproducibility of future studies.

**Methods:** A search of the PubMed database and a systematic manual screening of references until May 2024 for studies reporting peripheral bile acid concentrations in humans was conducted. English-language studies reporting mean or median concentrations of at least one of 15 predetermined circulating bile acids in human cohorts were included. The exclusion criteria were editorials, commentaries, letters to the editor, conference proceedings, abstracts, and monographs. Raw bile acid concentrations, subject demographics (number, average age, sex distribution, health status, fasted/fed status), the blood matrix analysed, the matrix volume analysed, the bile acid extraction process, and analytical technique when available were extracted by a single observer.

**Results:** 65 studies involving 215 cohorts were selected. Bile acid concentrations in normal cohorts exhibit large intervariability. The analytical technique used to measure bile acid concentrations, the fasted/fed status of patients at the time of sampling, the choice of blood collection matrix, the starting volume of this matrix, and the choice of protein precipitation solvent are found to be determinants of this variability.

**Limitations:** Only mean or median bile acid concentrations in study cohorts were extracted from studies and compared since bile acid concentrations are rarely reported in individual subjects. Analysing mean or median bile acid concentrations in study cohorts may not give a true sense of bile acid concentrations and therefore their determinants.

**Discussion:** Experimental, analytical and biological sources of mean peripheral bile acid concentration variability were identified. These must be standardised across future studies to clarify the potential of peripheral bile acids as biomarkers.

## Introduction

### Bile acids

Bile acids (BAs) comprise several molecules built on an identical sterol backbone. Cholic acid (CA) and chenodeoxycholic acid (CDCA) are primary BAs. They are metabolic products of cholesterol in liver hepatocytes (Fig 1). In humans, most BAs are conjugated to glycine (G-), with a minority conjugated to taurine (T-), in a ratio of 3:1 (G-:T-) on average (1). BA conjugation in hepatocytes is highly efficient and increases both their secretion into bile and their water solubility at acidic pH levels preventing precipitation in the duodenum (2), where BAs aid in the absorption of dietary fats.

**Fig 1.**
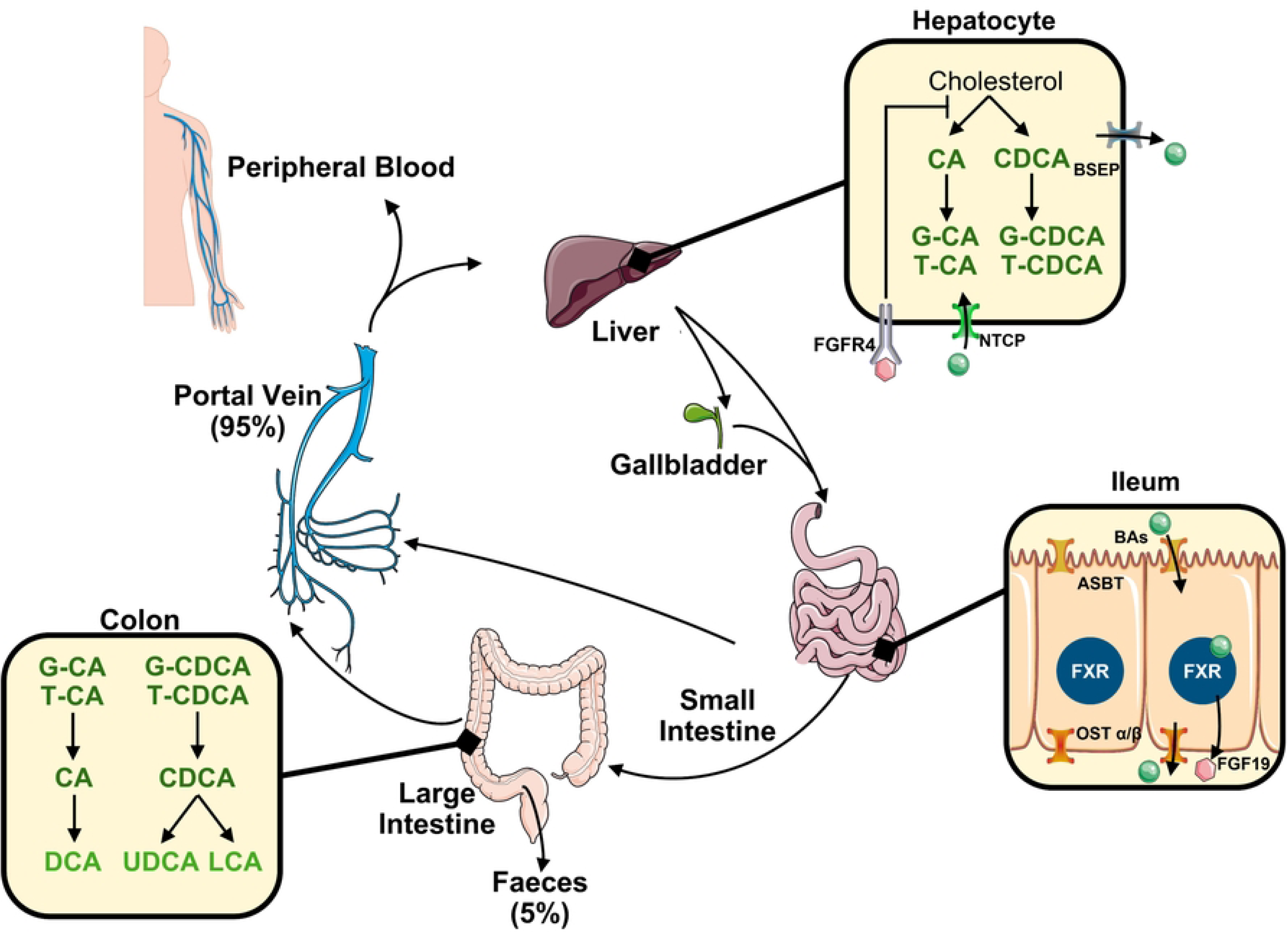
Schematic of the human enterohepatic circulation. Abbreviations: ASBT, Apical Sodium-Bile acid Transporter; BSEP, Bile Salt Export Pump; CA, cholic acid; CDCA, chenodeoxycholic acid; DCA, deoxycholic acid; FGF19, Fibroblast Growth Factor 19; FGFR4, Fibroblast Growth Factor Receptor 4; FXR, Farnesoid X Receptor; G-, glycine-conjugated; LCA, lithocholic acid; NTCP, Na^+^-Taurocholate Cotransporting Polypeptide; OST α/β, Organic Solute Transporter alpha/beta; T-, taurine-conjugated; UDCA, ursodeoxycholic acid.

From the liver, conjugated BAs (cBAs) are secreted through the bile salt export pump (BSEP; ABCB11) into the bile duct, from where they are stored in the gallbladder during periods of fasting, or flow directly into the duodenum during prandial periods. From the duodenum, BA flow continues through the jejunum and ileum. Throughout the small intestine, cBAs can be passively absorbed through the intestinal wall into the portal vein. However, the majority of cBA absorption occurs in the terminal ileum (3), where the apical sodium-bile acid transporter (ASBT; SLC10A2) mediates the active transport of BAs from the lumen of the ileum into enterocytes. In enterocytes, BAs bind to FXR, a nuclear receptor, with varying affinities according to BA species (4,5). Activation of FXR stimulates the production and secretion of fibroblast growth factor 19 (FGF19). The latter is brought to the liver with the portal flow where it binds to the FGFR4 on hepatocytes to inhibit the synthesis of BAs, functioning as an autoregulatory negative feedback mechanism. BAs are exported from enterocytes to the portal vein by the basolateral organic solute transporter (OST-α/β; SLC51A/SLC51B). BAs not actively transported into enterocytes of the terminal ileum continue their journey towards the colonic lumen where they are rapidly deconjugated by the gut microbial enzyme, bile salt hydrolase (BSH). The so formed unconjugated BAs (uBAs) can undergo dehydroxylation by cooperative bacterial enzymes, to form secondary BAs such as deoxycholic acid (DCA), lithocholic acid (LCA) and ursodeoxycholic acid (UDCA).

Lipophilic secondary uBAs are passively absorbed from the colonic lumen into colonocytes to the portal vein. Remaining colonic BAs are excreted in the faeces, accounting for approximately 5% of the BA pool daily. The portal vein returns BAs to the liver, where they are transported into hepatocytes by basolateral Na^+^-taurocholate cotransporting polypeptide (NTCP; SLC10A1), completing the enterohepatic circulation (EHC); while a minor fraction of BAs leak into the peripheral circulation.

### Bile acids as biomarkers

Besides their role in lipid digestion, BAs are important signalling molecules. BAs bind to multiple receptors, including FXR, Takeda-G-protein-receptor 5 (TGR5), pregnane X receptor (PXR), vitamin D receptor (VDR) and liver X receptor-α (6). The activation of these receptors contributes to the regulation of lipid homeostasis, glucose homeostasis, energy metabolism and inflammation (7). Each individual BA species binds to receptors with a specific affinity. Hence, the concentration of individual BAs at a given time and location along the EHC defines a specific activity which varies dynamically with time (for example in relation to a meal and release of massive amounts of BA in the gut) and position along the EHC. Many disease processes, ranging from cholestasis to digestive disorders and including dysmetabolic disorders, are associated with perturbed BA metabolism. Hence alteration of the size and composition of the BA pool, could be used as a biomarker of disease. Using peripheral BA concentrations as biomarkers has been investigated in various diseases such as obesity and insulin resistance (8–11), Type 2 Diabetes Mellitus (8,10,12–15), Metabolic Dysfunction-Associated Steatotic Liver Disease (MASLD) (9,16–29), cancers, such as hepatocellular carcinoma (30–33), cholangiocarcinoma (34) and colon cancer (35), and Alzheimer’s disease (36–38).

BAs could play a role in regulating both energy metabolism and inflammation, which is particularly relevant in the context of MASLD. In mice, the composition of the portal BA pool has been implicated in driving liver inflammation (39). Secondary BAs, produced through gut microbial metabolism are more potent activators of TGR5, a receptor found in the plasma membrane of Kupffer cells, the resident liver macrophages (40). TGR5 activation elevates cyclic adenosine monophosphate (cAMP), inhibiting the activation of nuclear factor kappa-light-chain-enhancer of activated B cells (NF-κB) (41), resulting in the inhibition of proinflammatory cytokine production. Consequently, a higher proportion of primary BAs relative to secondary BAs in the portal blood could exacerbate liver inflammation due to decreased TGR5 activation. Since sampling from the portal vein is extremely invasive, the question of using minimally invasive peripheral blood BA concentrations as a marker of MASLD severity might be raised. However, further research is needed to establish the relationship between peripheral BA levels and the composition of the portal BA pool, as well as their predictive value for MASLD severity. Of note, next to inflammation and metabolism, the activity of the thermogenic brown adipose, known to be defective in obesity and MASLD, is also in part modulated by BAs. Hence, BAs outside the EHC may also contribute to MASLD.

To date, all the existing studies on MASLD have only reported BA composition in the peripheral blood of MASLD patients. A recent systematic review found that total BA concentrations as well as circulating UDCA, T-CA, CDCA, T-CDCA, G-CA, G-UDCA, G-CDCA, T-UDCA and CA were significantly elevated (in order of most-elevated to least-elevated) in MASLD patients compared to healthy controls (42). Though the average concentrations of these BAs were increased, the intrinsic high inter-individual variability in BA concentrations (e.g. between subjects) meant that there was overlap between MASLD and healthy cohorts. But individual studies draw contradictory results. For example, Zhang et al. observed a decrease in peripheral G-DCA concentrations in MASLD patients compared to healthy controls (27); conversely Ferslew et al. observed an increase in peripheral G-DCA concentrations (29), while concentrations reported by Wu et al. exhibited no change (28).

Another review exploring the use of systemic BAs as biomarkers for diverse liver diseases showed that the studies did not agree on which individual BAs or BA ratios are relevant as biomarkers of specific liver dysfunction, including MASLD (43). Except for the prognostic value of systemic BAs in intrahepatic cholestasis of pregnancy, studies have failed to provide evidence for using systemic BAs as a liver biomarker. This study highlighted inconsistencies in the reported results related, in part, to differences in the methods used in BA assays.

### Metadata on bile acid data

Contextualizing published data on BA concentrations is critical to validate their potential use as biomarkers. However, most studies are conducted on small sample sizes making it difficult to extrapolate results to a whole population. Additionally, study conclusions are often inconsistent, highlighting the need to better understand the potential causes of these discrepancies. To address this, we propose a systematic review of the literature to investigate the influence of biological determinants, such as health status, type of disease, age and fed status, as well as experimental factors on BA data reporting.

## Methods

### Literature search

The literature search was performed in PubMed as of May 2024 using the following search: “bile acid concentration”[Title/Abstract] OR “bile acid concentrations”[Title/Abstract] OR “circulating bile acid”[Title/Abstract] OR “circulating bile acids”[Title/Abstract] OR “bile acid profile”[Title/Abstract] OR “bile acid profiles” OR “plasma bile acid”[Title/Abstract] OR “plasma bile acids”[Title/Abstract] OR “serum bile acid”[Title/Abstract] OR “serum bile acids”[Title/Abstract] OR “individual bile acids “[Title/Abstract] OR “bile acid levels”[Title/Abstract]. The review and the review protocol were not registered. The search was complemented by a manual screening of the list of references for relevant articles based on predetermined inclusion and exclusion criteria.

### Inclusion and exclusion criteria

Studies that reported absolute peripheral concentrations of one or more of the 15 individual BAs or absolute peripheral concentrations of total BAs were included. Only studies written in English and conducted on humans were included. Editorials, commentaries, letters to the editor, conference proceedings, abstracts, and monographs were not included. These were determined by a single observer.

### Data and metadata extraction

The data for the most abundant primary and secondary conjugated and unconjugated bile acids (CA (PubChem CID:221493), CDCA (PubChem CID:10133), DCA (PubChem CID:222528), UDCA (PubChem CID:31401), LCA (PubChem CID:9903), G-CA (PubChem CID:10140), G-CDCA (PubChem CID:12544), G-DCA (PubChem CID:3035026), G-UDCA (PubChem CID:12310288), G-LCA (PubChem CID:115245), T-CA (PubChem CID:6675), T-CDCA (PubChem CID:387316), T-DCA (PubChem CID:2733768), T-UDCA (PubChem CID:9848818), T-LCA (PubChem CID:439763)) were extracted directly from the main publication or supplementary material of studies. If the arithmetic mean concentration was not reported, the geometric mean or median was extracted and presented as the arithmetic mean in this study. In cases where data were only presented in graphical form and not in tabular form, the graphs were digitised using WebPlotDigitizer (automeris.io) when possible. When data for the individual BA concentrations were not available, the total BA data were reported (totalCA, totalCDCA, totalDCA, totalUDCA, totalLCA). When the data for individual BA concentrations were available, but the total BA concentrations were not, concentrations of the conjugated and unconjugated forms of BAs were summed e.g. totalCA = CA + G-CA +T-CA. If BA concentrations of patients at multiple postprandial timepoints were available, these were also included as a separate set of data. The units of concentration were standardised to nM for comparison, using molecular weights reported in PubChem (https://pubchem.ncbi.nlm.nih.gov) where necessary. These data were extracted by a single observer.

Regarding the metadata, biological information was retrieved including the health status of the subjects, whether the subjects were in a fasted or fed state, the average age of the cohorts and the size and sex distribution of the cohorts when available. A cohort of patients was considered *unhealthy* if they had a condition that could directly impact the EHC. Health conditions included: cirrhosis, obstructive jaundice, (mild and severe) cholestasis, cholelithiasis, adenomyoma, ileal resection, primary sclerosing cholangitis, non-alcoholic fatty liver (NAFL), non-alcoholic steatohepatitis (NASH), hepatocellular carcinoma (HCC), cholangiocarcinoma, gallbladder cancer, primary colon cancer, viral hepatitis B and C, alcoholic liver disease, and various combinations thereof. By contract, we defined *normal* individuals here as ‘having no known diseases directly affecting the organs involved in the EHC’.

Then, the material used for analyses was recorded (serum or plasma). Additionally, the method by which BAs were extracted from this material and the analytical technique used to quantify the BA concentrations was extracted. For the analytical technique, there are many forms of Liquid Chromatography-Mass Spectrometry (LC-MS). Studies using any of these forms were clumped together and identified as using LC-MS in this study. In the studies utilising LC-MS, information about whether the sample was directly injected or whether steps were taken to extract BAs was obtained. This included information concerning what reagent was used for protein precipitation, whether the resulting mixture underwent solid phase extraction (SPE), whether the resulting mixture sample was evaporated, if so, was the sample evaporated under vacuum or nitrogen. And once evaporated, what reagent was used to reconstitute the residue before being injected into the LC-MS equipment. Missing or unclear information was reported as such.

### Visual plotting and statistics

The data were plotted using Python (Version 3.12.4) and graphed as individual points and presented as the mean ± standard deviation. Statistical analyses were performed using a combination of GraphPad Prism 8 and the SciPy Statistical functions (Version 1.14.0).

## Results

### Literature search and relevant studies

4,172 studies were returned following a search of the keywords (Fig 2). After applying the exclusion criteria and reading their abstracts and results sections a total of 65 studies reporting peripheral bile acid concentrations were included. Initially there was one additional study, but this was excluded because the BA concentrations reported appeared to be 1000-times higher than those in the other studies (44). There was no response when both the lead and corresponding authors were contacted to confirm whether the unit used was correct and hence the study was excluded. For another study the authors did respond and confirmed that the unit reported was incorrect (21). The 65 studies were published between 1977 and 2024. The studies included a total of 10629 individuals, of which 3249 were considered *normal* individuals, who we defined here as ‘having no known enterohepatic disorders or diseases directly affecting the organs involved in the EHC’ and 4628 were considered *unhealthy*, who we defined here as ‘having diagnosed enterohepatic disorders’. Over all studies, 3 studies which involved 471 individuals had both normal and unhealthy individuals in the same cohorts, while 6 studies did not report whether individuals had enterohepatic disorders and were thus classified as having *unknown health statuses*. From these 65 studies, BA concentration datasets from a total of 215 cohorts were obtained. 169 (∼79%) of these datasets reported individuals BAs (Table 1), with the remaining 46 (∼21%) reporting total BAs (Table 2). 101 (∼51%) were obtained from normal cohorts and 98 (∼46%) from unhealthy cohorts. 133 (∼62%) were from fasted cohorts and 59 (∼27%) were from fed cohorts. The material analysed was serum samples for 158 (∼73%) and plasma samples for 57 (∼27%). Regarding analytical technique, 154 (∼72%) were measured using LC-MS, 43 (20%) using Gas Chromatography Mass Spectrometry (GC-MS), 8 (∼4%) using Gas Liquid Chromatography (GLC), 3 (∼1%) using High Performance Liquid Chromatography (HPLC) and 4 (∼2%) using Capillary Electrophoresis.

**Fig 2.**
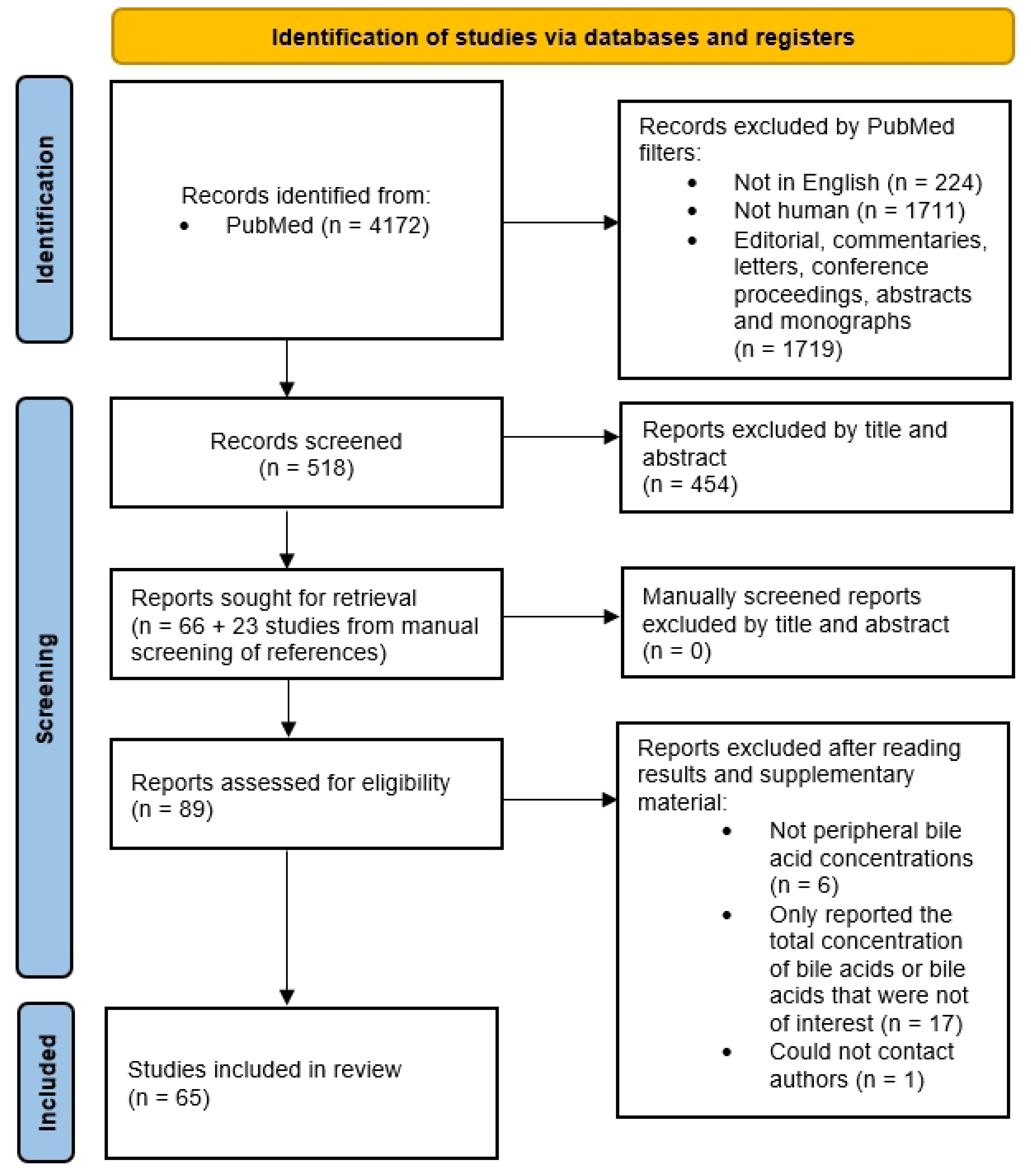
The flowchart based on PRISMA standards.

**Table 1.**
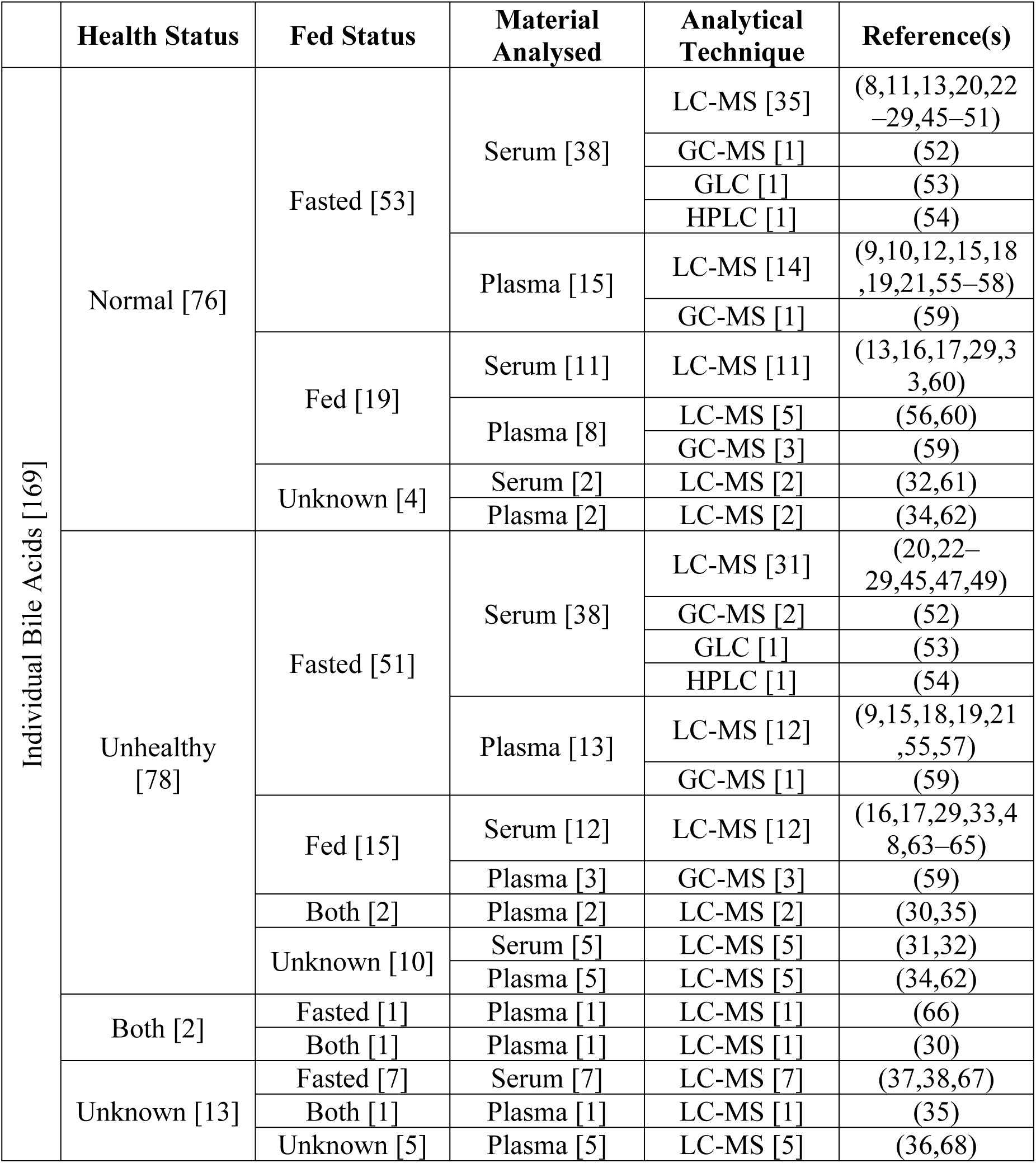
Biological and experimental characteristics of the individual bile acid concentration datasets obtained from the included studies. The number in squared brackets indicates the number of datasets in each subgroup. Abbreviations: LC-MS, liquid chromatography-mass spectrometry; GC-MS, gas chromatography-mass spectrometry; GLC, gas liquid chromatography; HPLC, high performance liquid chromatography.

**Table 2.** Biological and experimental characteristics of the total bile acid concentration datasets obtained from included studies. The number in squared brackets indicated the number of datasets in each subgroup. Abbreviations: LC-MS, liquid chromatography-mass spectrometry; GC-MS, gas chromatography-mass spectrometry; GLC, gas liquid chromatography; HPLC, high performance liquid chromatography.

|  | Health Status | Fed Status | Material Analysed | Analytical Technique | Reference(s) |
| --- | --- | --- | --- | --- | --- |
| Total Bile Acids [46] | Normal [25] | Fasted [7] | Serum [7] | GC-MS [4] | (65,69–71) |
|  |  |  |  | GLC [2] | (63,72) |
|  |  |  |  | Capillary Electrophoresis [1] | (73) |
|  |  | Fed [18] | Serum [18] | LC-MS [1] | (17) |
|  |  |  |  | GC-MS [16] | (65,69,70) |
|  |  |  |  | GLC [1] | (63) |
|  | Unhealthy [20] | Fasted [13] | Serum [13] | GC-MS [7] | (65,71,74–77) |
|  |  |  |  | GLC [2] | (63,72) |
|  |  |  |  | HPLC [1] | (78) |
|  |  |  |  | Capillary Electrophoresis [3] | (73) |
|  |  | Fed [7] | Serum [7] | LC-MS [1] | (17) |
|  |  |  |  | GC-MS [5] | (65) |
|  |  |  |  | GLC [1] | (63) |
|  | Both [1] | Fasted [1] | Plasma [1] | LC-MS [1] | (14) |

### Normal cohorts exhibit large intervariability in peripheral BA concentrations

The concentrations of individual BAs in normal cohorts are highly variable between cohorts for a given BA (Fig 3A). In general, CA, CDCA and DCA in their free and conjugated forms are the most abundant compared to these forms of UDCA and LCA. G-CDCA is the most abundant BA, with concentrations ranging between 30nM and 2000nM. This is followed by CDCA and DCA, which both have similar mean concentrations. In contrast, T-LCA and T-UDCA are the least abundant BAs, with mean concentrations ∼10nM. The mean concentrations of G-conjugated BAs are between two- and ten-fold higher than their T-conjugated counterparts.

**Fig 3.**
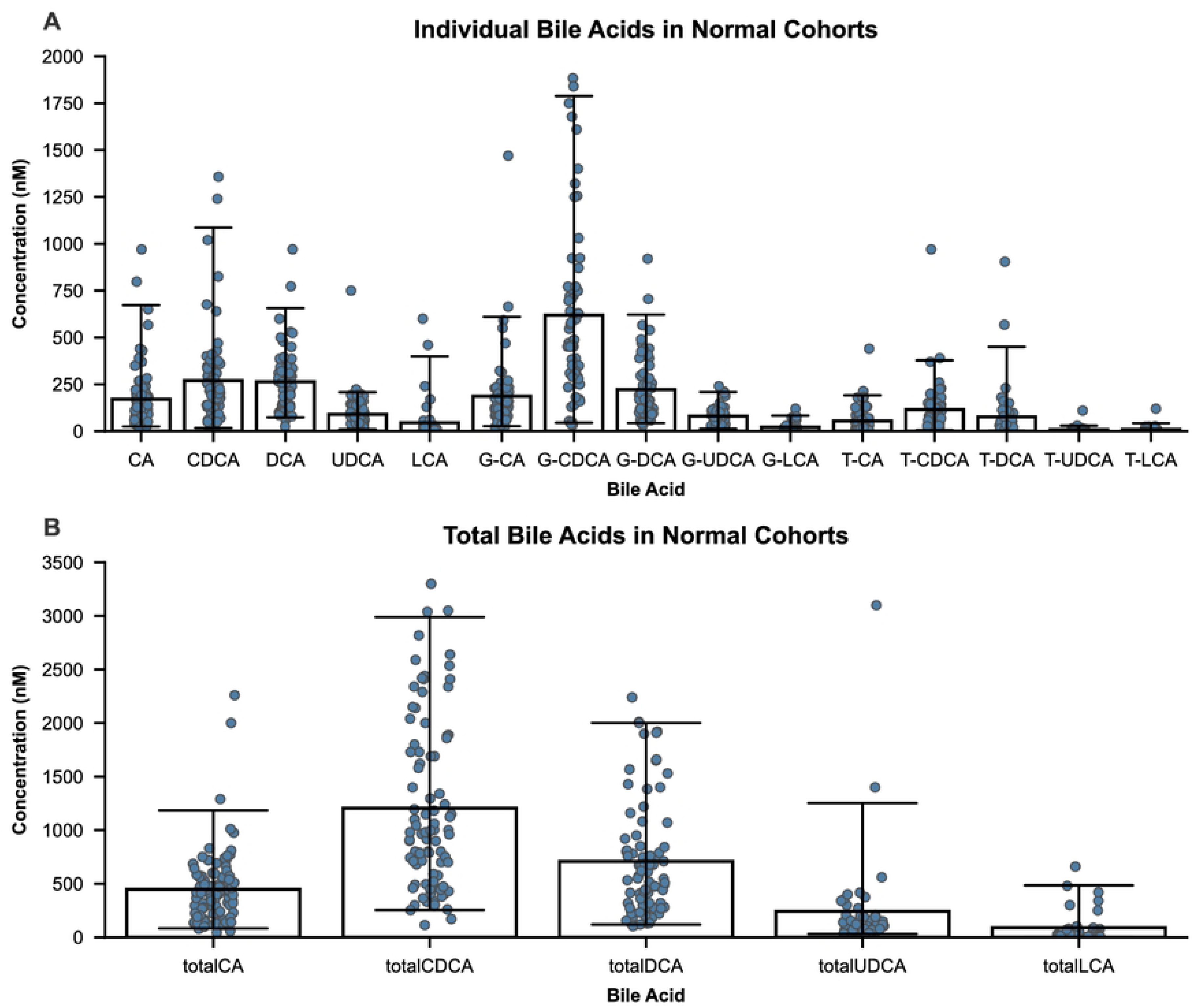
Peripheral bile acid concentrations in normal cohorts are highly variable. Mean/Median individual bile acid concentrations (A) and total bile acid concentrations (B) in normal cohorts (those without enterohepatic disorders) were collected from literature and plotted as their mean ± standard deviation.

The concentration of total BAs also fluctuates greatly between cohorts (Fig 3B), with totalCDCA being the most abundant. This is followed by totalDCA, totalCA, totalUDCA and totalLCA in descending order. totalCDCA has the widest range from 100nM to 1900nM, while totalLCA is the smallest, ranging from 0nM to 700nM.

### LC-MS is currently the most used analytical technique to measure circulating individual BA concentrations

In the past, HPLC and GLC were used to quantify BA concentrations. However, owing to the higher sensitivity, specificity and throughput of LC-MS and GC-MS, HPLC and GLC are now obsolete. Consequently, the vast majority of studies in which BA concentrations are reported have used either LC-MS or GC-MS (Fig 4). Of the 65 studies included here, 49 studies used LC-MS and 10 studies used GC-MS. The fact that GC-MS requires a complex, time-consuming derivatisation step to convert the BAs to a volatile form, and that LC-MS has a higher sensitivity and throughput, has led to LC-MS becoming the primary analytical technique for measuring individual BAs (Table 1), while studies in which GC-MS was used reported total BAs (Table 2).

**Fig 4.**
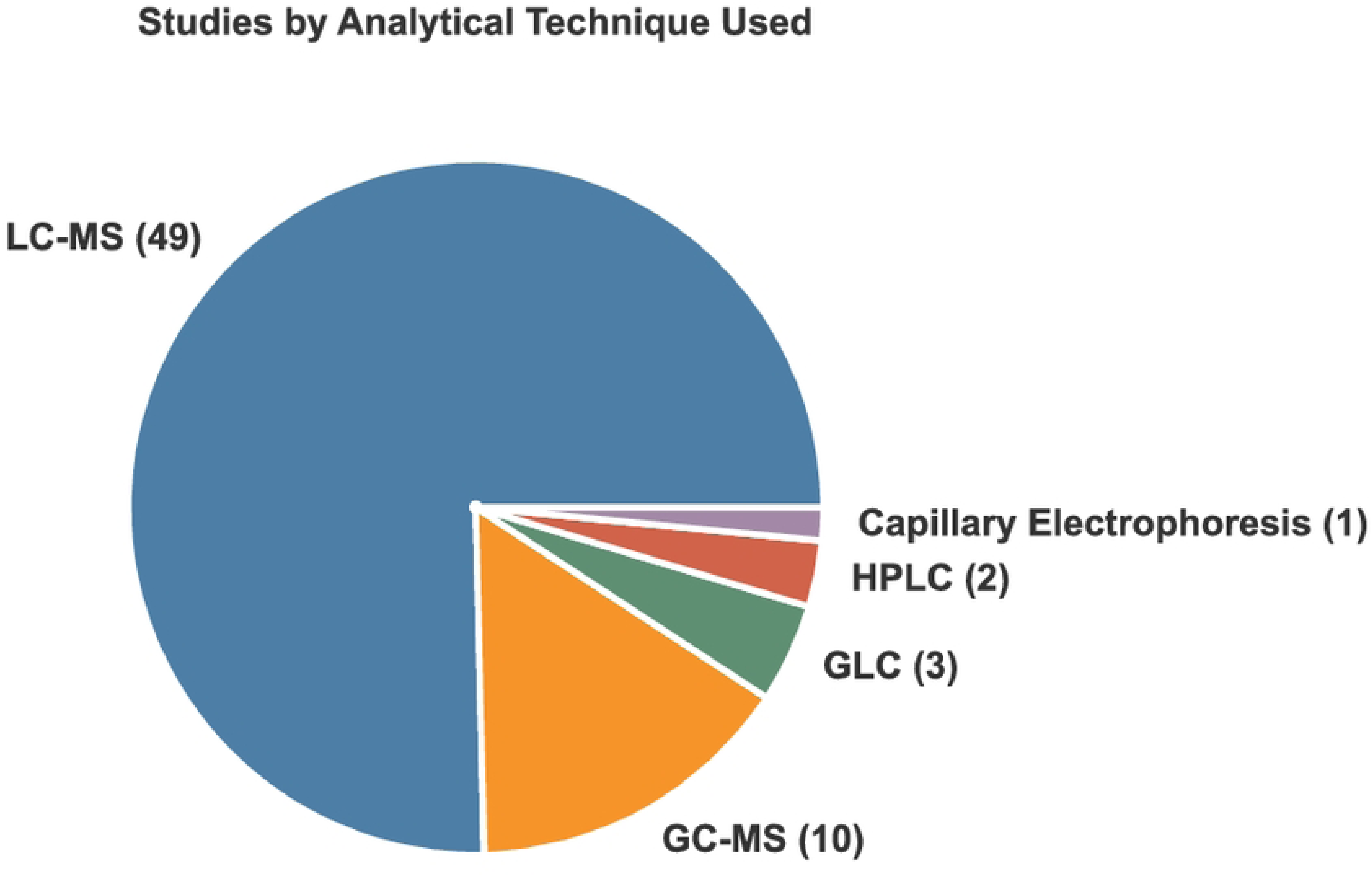
LC-MS is the predominant analytical technique to measure bile acid concentrations. The analytical technique used in the 65 included studies were tallied and presented as a pie chart (A). The number in brackets indicates the number of studies using the respective technique. Abbreviations: GC-MS, gas chromatography-mass spectrometry; GLC, gas liquid chromatography; HPLC, high performance liquid chromatography.

### The fed status at the time of sampling has an impact on BA concentrations

The impact of the fasted/fed status at the time of sampling on the concentration of individual BAs in normal cohorts was investigated. Having established that LC-MS is the most widely used analytical technique to measure BA concentrations, we restricted our analysis to LC-MS studies. Normal cohorts fed at the time of sampling tend to have higher mean concentrations of G-BAs than fasted cohorts (Fig 5A). The mean concentrations of G-CDCA (p=0.0396), G-DCA (p=0.00007), G-UDCA (p=0.0111) and G-LCA (p=0.0016) are increased, but not significantly for mean concentrations of G-CA. Of the uBAs, mean DCA concentrations are significantly higher in fed normal cohorts (p=0.0061), while mean concentrations of CA, CDCA, UDCA and LCA are similar in both conditions. T-CDCA is the only T-BA that is significantly elevated in fed normal cohorts (p=0.0081), while the other T-BAs remain similar in both conditions. The totalDCA mean concentrations are increased in fed normal cohorts compared to fasted (p=0.00091), while the mean concentrations of the other totalBAs remain similar in both conditions, though data for totalUDCA in fed normal cohorts are limited (n=3) (Fig 5B).

**Fig 5.**
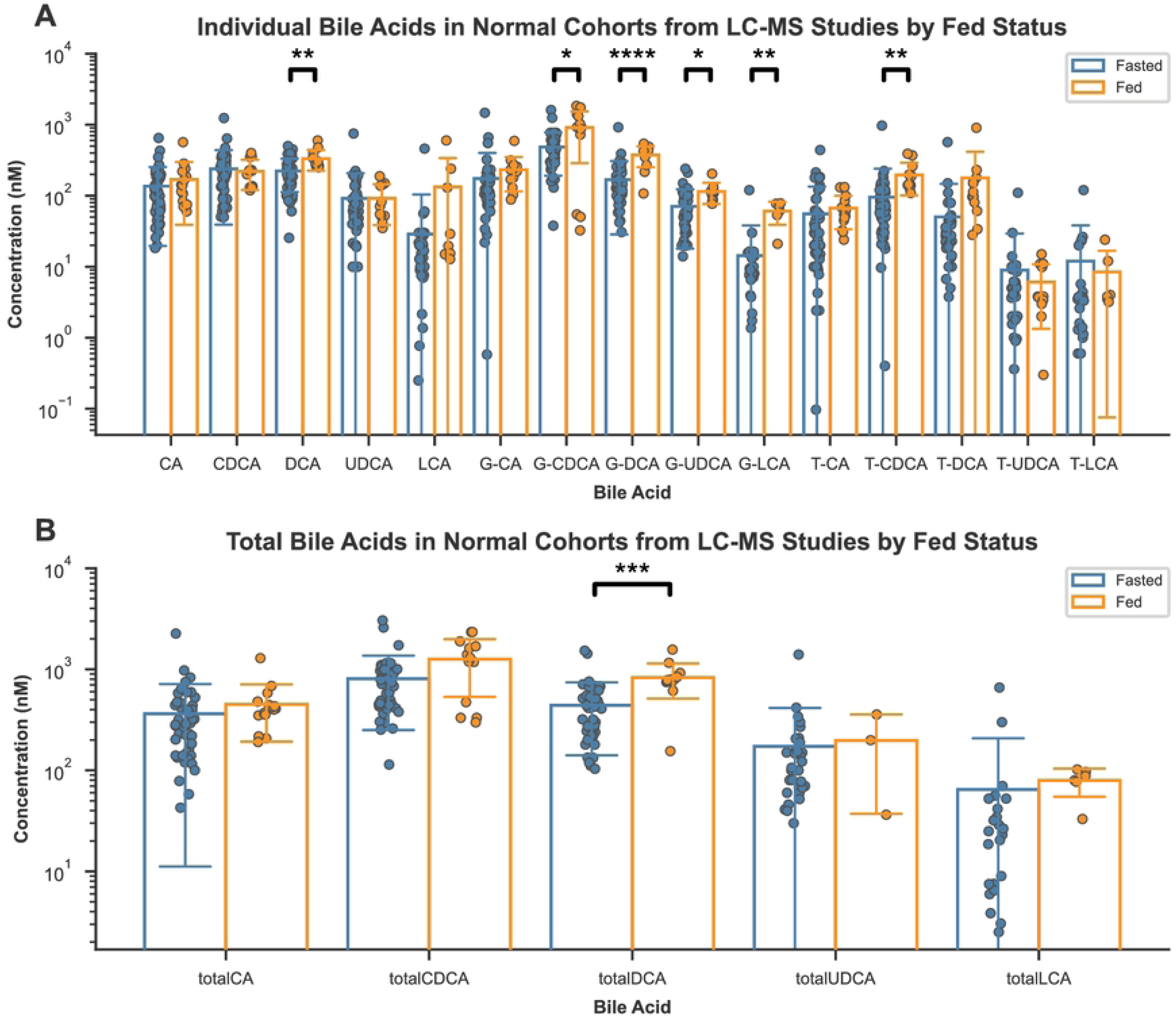
Normal cohorts fed at the time of sampling tend to have elevated mean bile acid concentrations than fasted normal cohorts. Mean/median individual bile acid (A) and total bile acid (B) concentrations from studies that used LC-MS in normal cohorts (those without enterohepatic disorders) were stratified for whether the cohorts were fasted or fed at the time of sampling and plotted as their mean ± standard deviation. Statistics performed by two-tailed unpaired Welch’s t-test, blank p>0.05, * p≤0.05, ** p≤0.01, *** p≤0.001, **** p≤0.0001.

Since the fasted/fed status at the time of sampling has an impact on BA concentrations, reducing the variability associated with it might be relevant. Due to *nil per os* guidelines for preoperative patients, samples for BA analyses should always be obtained in the fasted state, to minimise any variation caused by a subject’s fasted/fed status at the time of sampling, unless specifically investigating postprandial effects.

### BA concentrations in peripheral serum are higher than peripheral plasma

Peripheral blood can be collected as either serum or plasma. To collect serum, blood is allowed to clot naturally and then centrifuged, enabling the removal of coagulation factors, blood cells and fibrin clots. On the other hand, plasma is collected by first adding an anticoagulant such as heparin, EDTA or citrate, and then immediately centrifuged to separate blood cells and liquid plasma.

Serum samples obtained from normal cohorts fasted at the time of sampling tend to have higher individual BA concentrations than plasma samples on average (Fig 6A). The mean concentrations of CA (p=0.0064), CDCA (p=0.0051), UDCA (p=0.0271), G-CA (p=0.0477) and T-CA (0.0090) in serum samples are significantly higher than those in plasma. The mean concentrations of totalBAs are also similar in both serum and plasma samples, with only totalCA significantly higher in serum samples (p=0.0133) (Fig 6B). Though serum samples exhibited higher BA concentrations on average, across both individual BAs and total BAs, plasma samples seem to exhibit lower variation compared to serum samples, suggesting that BA concentrations in plasma may be more reliable and reproducible. However, a comparison of variance via a *Brown-Forsythe test* reveals no statistically significant difference between the variance of two groups across all BAs (S3 Table).

**Fig 6.**
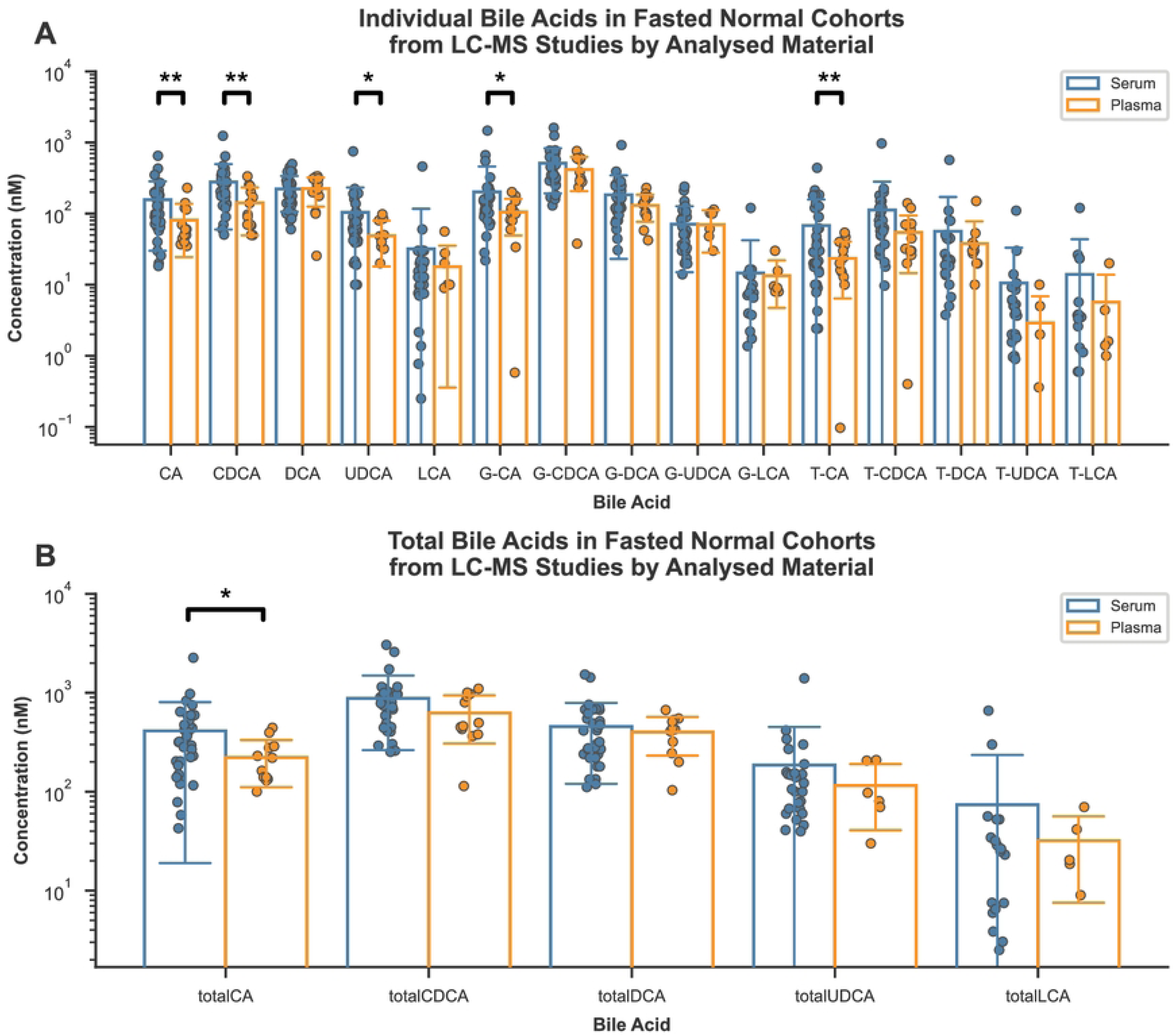
Bile acid concentrations in peripheral serum tend to be higher than those in peripheral plasma. Mean/median individual bile acid (A) and total bile acid (B) concentrations from studies that used LC-MS in normal cohorts (those without enterohepatic disorders) which were fasted at the time of sampling were stratified for whether the sample analysed was serum or plasma, and plotted as their mean ± standard deviation. Statistics performed by two-tailed unpaired Welch’s t-test, blank p>0.05, * p≤0.05, ** p≤0.01.

### A wide range of bile acid concentrations can be detected from a 50µl plasma sample

The sensitivity of the LC-MS instrumentation can play a role in the variability of concentrations measured. If the starting volume is too small, the amount of a BA in this volume, particularly those in low concentration might fall below the detection limit of the instrument.

Of the 14 datasets of BA concentrations measured using LC-MS in plasma from normal cohorts fasted at the time of sampling, half of them were measured in 50µl of plasma (Fig 7A). The next most used volume of plasma was 100µl. Similar ranges of total BA concentrations can be measured from both 50µl and 100µl of plasma (Fig 7B). This is also true for G-CDCA, the most abundant BA of the 15 BAs focussed on in this study, while there are no measurements of T-UDCA, the least abundant BA of those focussed on here, from 100µl of plasma (S4 Fig). No correlation between the starting volume of plasma and the concentration of BAs measured is observed. Thus, 50µl of plasma appears to be a suitable volume to use to ensure accurate absolute BA concentration measurements whilst potentially reducing the volume of blood required to be drawn from a subject.

**Fig 7.**
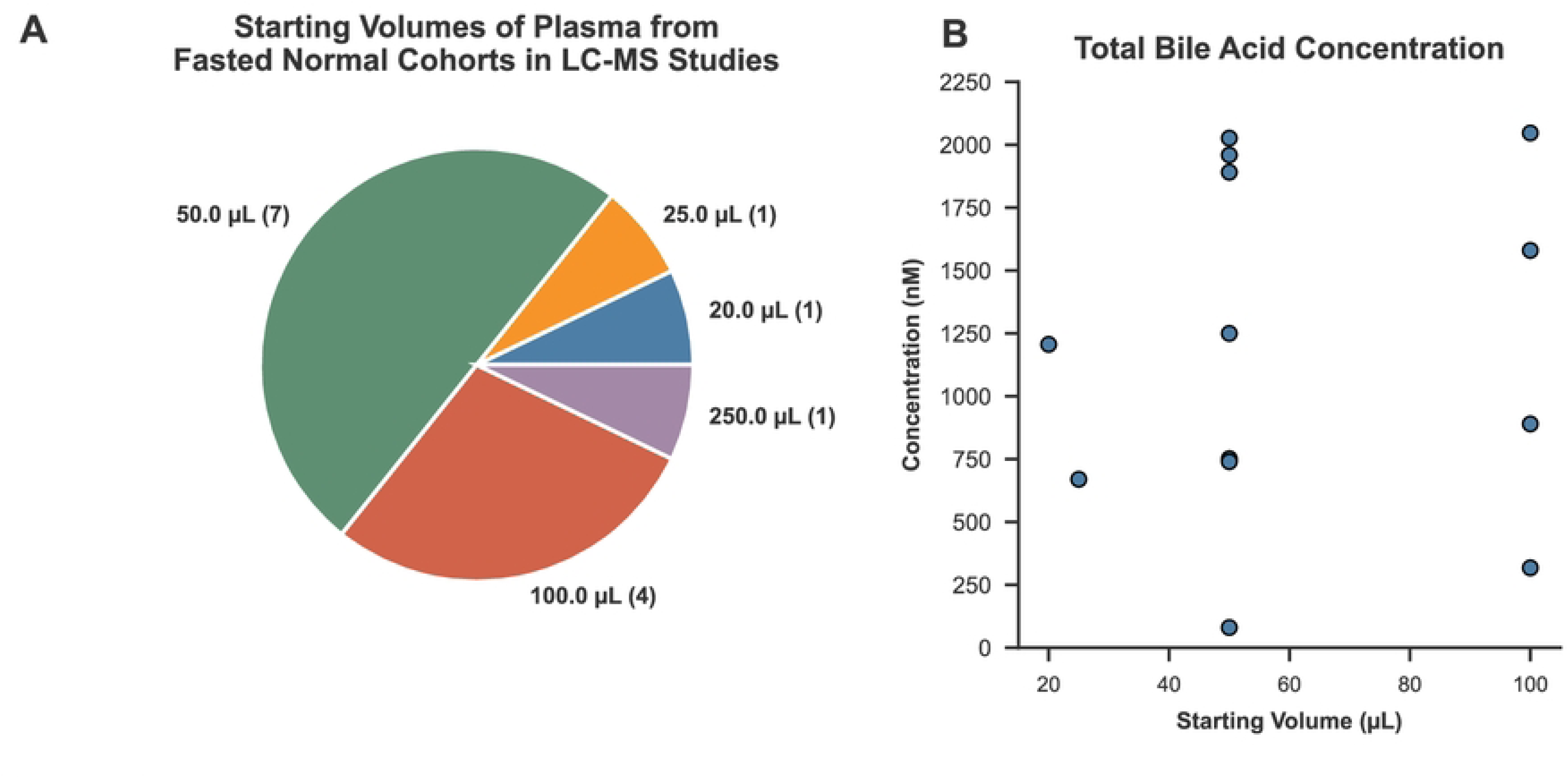
Peripheral bile acid concentrations are most commonly measured in 50µl of plasma and a wide range of concentrations can be detected. The starting volume of plasma used to measure bile acid concentrations from studies that used LC-MS in normal cohorts (those without enterohepatic disorders) which were fasted at the time of sampling were tallied and presented as a pie chart (A). The number in brackets indicates the number of datasets with the respective starting volume of plasma. The total bile acid concentration of each dataset was summed and plotted versus their starting volumes of plasma (B).

### When methanol is used to precipitate proteins, a wider range of BA concentrations can be measured across all BAs

A critical step in the upstream processing of a sample to be analysed by LC-MS is protein precipitation, during which proteins in serum and plasma that would otherwise interfere with BA analysis by binding to BAs or clogging the analytical column, are removed. The two most common methods involve using either an organic solvent for precipitation, such as methanol, ethanol or acetonitrile, or an acid for precipitation, such as formic acid or trichloroacetic acid. Another method is alkaline hydrolysis which involves the addition of an alkaline such as sodium hydroxide, followed by heating to facilitate hydrolysis and then centrifugation to remove the precipitated proteins. However, exposure to strong alkaline conditions may lead to the degradation of BAs.

Of the seven BA concentration datasets measured in 50µl of plasma from normal cohorts in the fasted state using LC-MS, methanol was used to precipitate proteins in four of them, acetonitrile, two, and sodium hydroxide, one (Fig 8A). The total BA concentrations in samples in which proteins were precipitated with methanol are similar to those precipitated with acetonitrile (Fig 8B). For G-CDCA, its concentrations are similar in both cases (Fig 8C), while concentrations of T-UDCA are higher when methanol is used compared to when acetonitrile is used (Fig 8D). When concentrations for 100µl of plasma are included, since no difference between using 50µl and 100µl of plasma was observed in total BA concentrations measured (Fig. 7B), the concentrations of G-conjugated and T-conjugated CA, CDCA and DCA are similar when methanol and acetonitrile are used, but concentrations of G-conjugated and T-conjugated UDCA and LCA are larger when methanol is used, though data are limited (S5 Fig).

**Fig 8.**
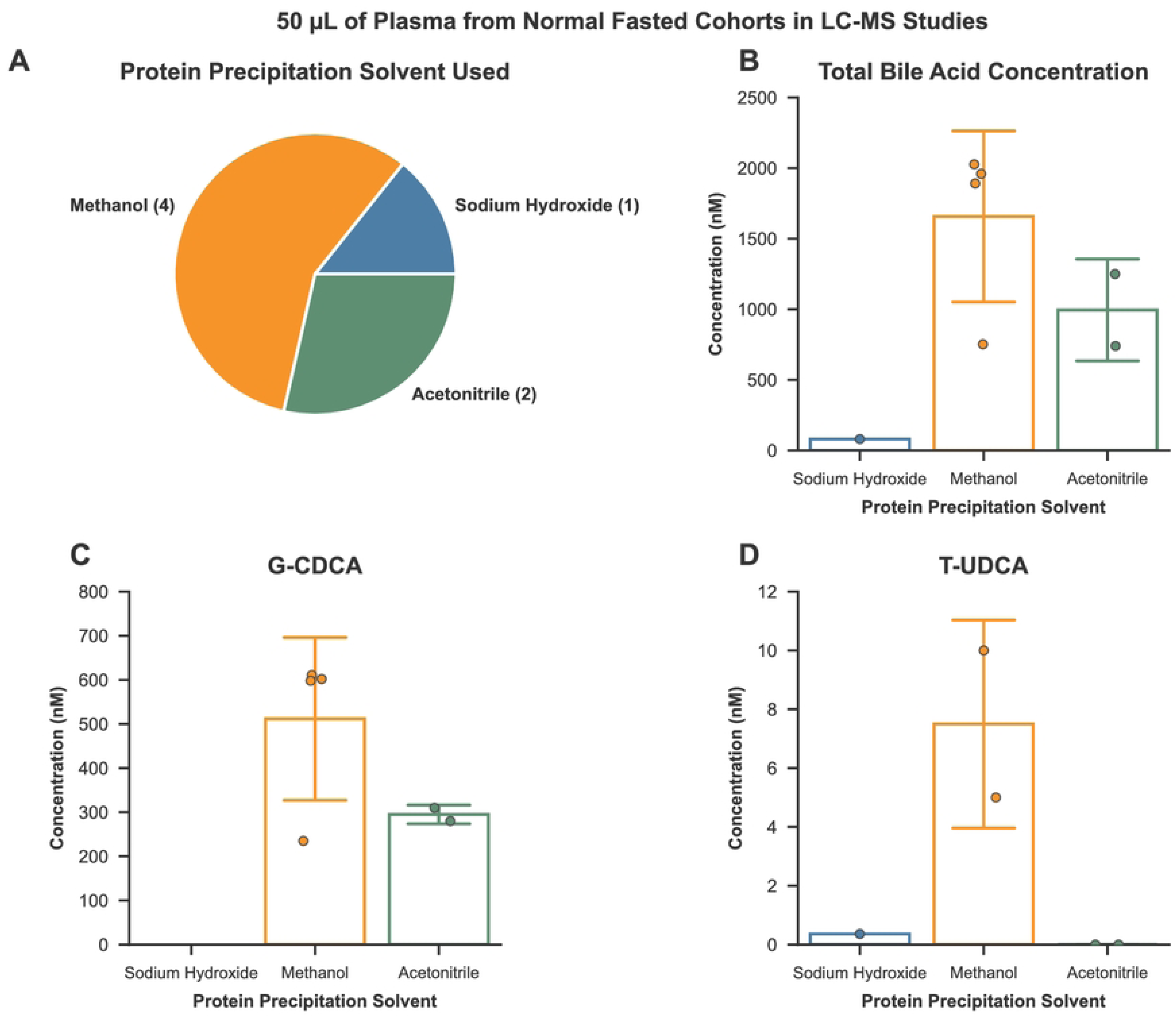
Bile acid concentrations in plasma samples in which proteins were precipitated with methanol tend to be higher across all bile acids than when other solvents are used. The protein precipitation solvent used in datasets from studies that used LC-MS to measure bile acid concentrations in 50µL plasma from normal cohorts (those without enterohepatic disorders) which were fasted at the time of sampling were tallied and presented as a pie chart (A). The total bile acid concentrations of these datasets were summed and stratified for the protein precipitation solvent used and plotted as their mean ± standard deviation (B). The concentrations of the most abundant bile acid: G-CDCA (C) and the least abundant bile acid: T-UDCA (D) were stratified for the protein precipitation solvent used and plotted as their mean ± standard deviation.

Having identified determinants contributing to explain the variability observed in reported BA concentrations in human peripheral blood, we aimed to see if, by filtering studies for the suggested recommendations (Table 3), we could reduce the high intervariability observed in BA concentrations in normal cohorts (those without enterohepatic disorders). A comparison of variance via a *Brown-Forsythe test* found that G-UDCA (p=0.04925) is the only BA whose variance is significantly decreased when the data is filtered (S6 Fig and S7 Table).

**Table 3.** The identified sources of variability in bile acid concentration measurements and the recommendations to reduce this variability. Abbreviations: LC-MS, liquid-chromatography-mass spectrometry.

| Source of Variability | Recommendation |
| --- | --- |
| Analytical Technique | LC-MS |
| Fasted/Fed Status of Subjects | Fasted |
| Analyzed Material | Plasma |
| Starting Volume | 50μl |
| Protein Precipitation Solvent | Methanol |

### Unhealthy cohorts exhibit increased BA concentrations compared to normal cohorts

Without filtering for the suggested recommendations, unhealthy cohorts exhibit increased mean individual BA concentrations across most BAs compared to normal cohorts (Fig 9A). Concentrations of UDCA (p=0.0205), G-CA (p=0.0041), G-CDCA (p=0.0002), G-DCA (p=0.0014), T-CA (p=0.0144), T-CDCA (p=0.0022), T-DCA (p=0.0233) and T-UDCA (p=0.0338) are significantly elevated in unhealthy cohorts, while concentrations of the other BAs are similar in both groups of cohorts on average. totalCA (p=0.0024), totalCDCA (p=0.00002), totalDCA (p=0.0066), totalUDCA (p=0.0238) are significantly elevated in unhealthy cohorts, while totalLCA remains similar in both normal and unhealthy cohorts (Fig 9B).

**Fig 9.**
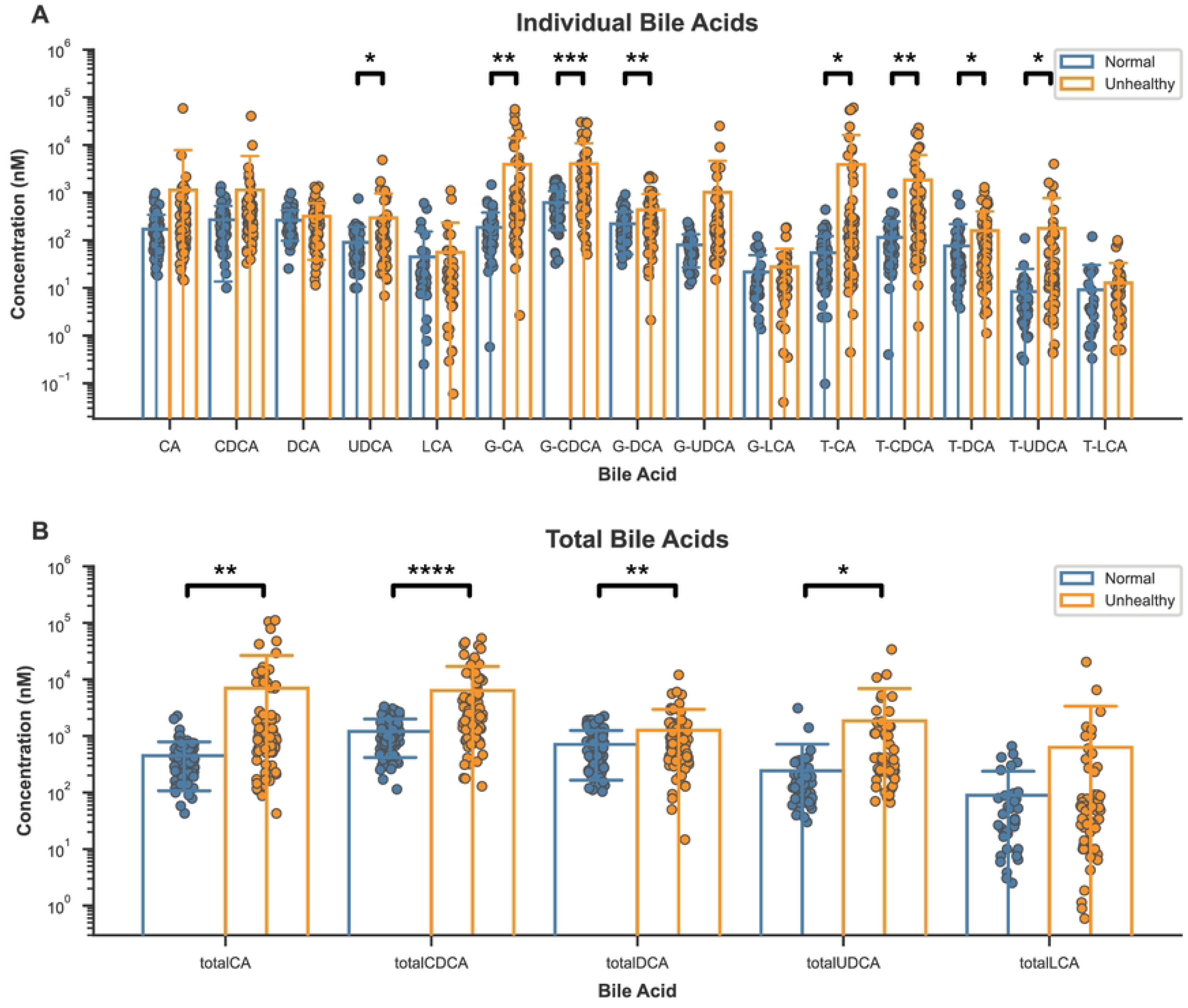
Unhealthy cohorts (those with enterohepatic disorders) exhibit elevated bile acid concentrations compared to normal cohorts (those without enterohepatic disorders). Mean/median individual bile acid (A) and total bile acid (B) concentrations in normal (without enterohepatic disorders) and unhealthy cohorts (with enterohepatic disorders) were plotted as their mean ± standard deviation. Statistics performed by two-tailed unpaired Welch’s t-test, blank p>0.05, * p≤0.05, ** p≤0.01, *** p≤0.001, **** p≤0.0001.

Diseases were stratified into three groups: NAFLD+Cirrhosis, HCC and Cholestasis. Cohorts with known instances of NAFL, NASH and Cirrhosis were placed in the NAFLD+Cirrhosis group, while cohorts with known instances of HCC or Cholestasis were placed in the HCC or Cholestasis groups respectively. When stratified for diseases, there is a significant difference in the total BA concentration between normal cohorts and cholestatic cohorts (p=0.0399) (Fig 10A), in line with the literature. However, there is no difference between cholestatic cohorts and cohorts with metabolic-dysfunction associated liver diseases (NAFLD+Cirrhosis) and HCC. There is a significant difference between the conjugated-to-unconjugated BA ratios in HCC cohorts and those in normal (p=0.0007) and NAFLD+Cirrhotic cohorts (p=0.0003) (Fig 10B). The secondary-to-primary BA ratios in both HCC and cholestatic cohorts are significantly lower than the secondary-to-primary BA ratios in normal (p=0.00001, p=0.0006 respectively) and NAFLD+Cirrhotic cohorts (p=0.00009, p=0.0022 respectively) (Fig 10C).

**Fig 10.**
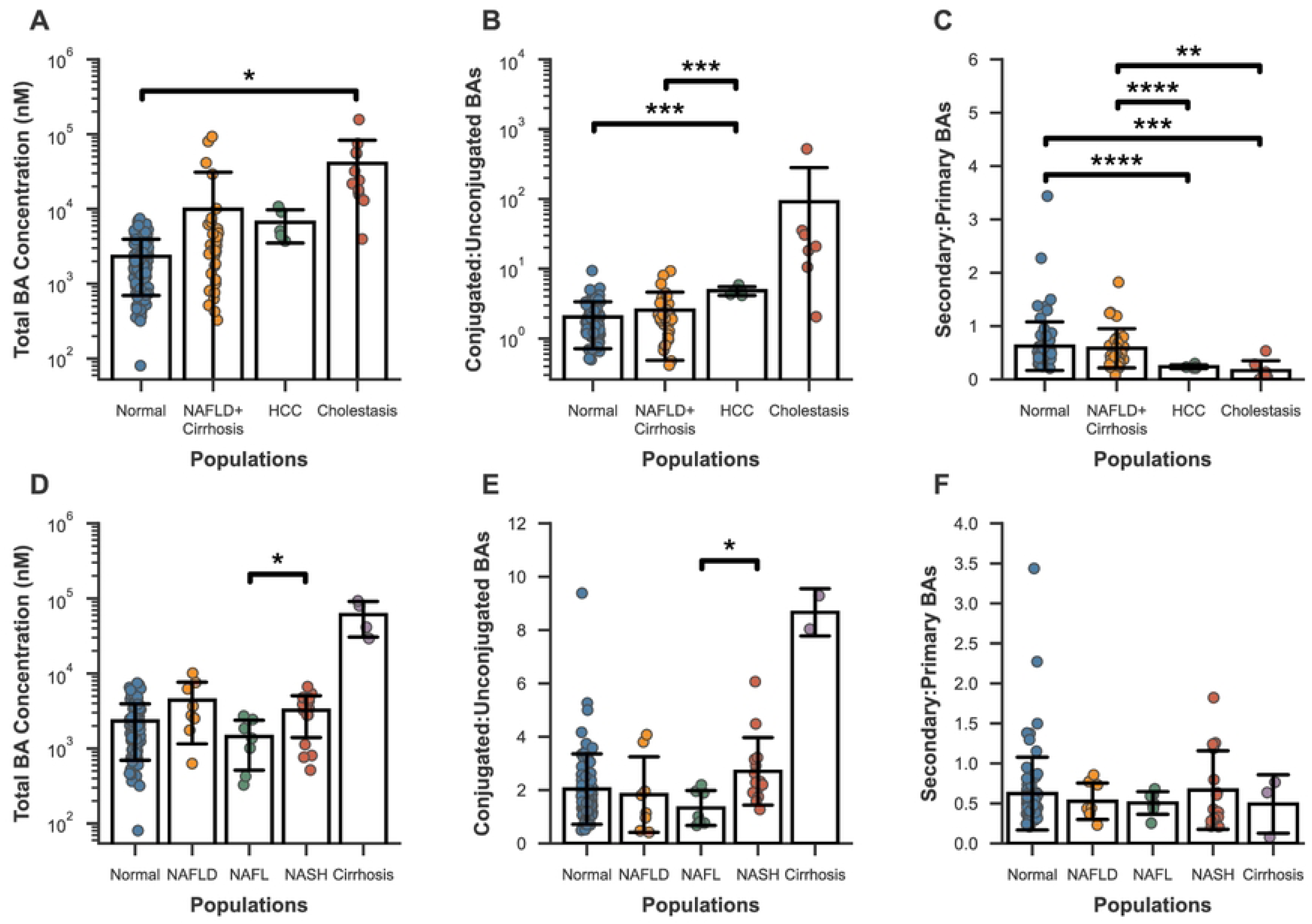
Circulating bile acids are a poor marker of liver diseases. The mean/median of the total bile acid concentrations (A), conjugated-to-unconjugated bile acid ratios (B) and secondary-to-primary bile acid ratios (C) in normal cohorts (those without enterohepatic disorders), cohorts with metabolic-dysfunction associated liver disorders (NAFLD + cirrhotic), HCC, and cholestatic cohorts were plotted as their mean ± standard deviation. The mean/median of the total bile acid concentrations (D), conjugated-to-unconjugated bile acid ratios (E) and secondary-to-primary bile acid ratios (F) in normal, NAFLD, NAFL, NASH and cirrhotic populations were plotted as their mean ± standard deviation. Statistics performed by Games-Howell Multiple Comparison Test, blank p>0.05, * p≤0.05, ** p≤0.01, *** p≤0.001, **** p≤0.0001.

Categorising the NAFLD+Cirrhotic cohorts by the stage of NAFLD progression into NAFL (non-alcoholic fatty liver), NASH (non-alcoholic steatohepatitis) or cirrhosis shows that cohorts with NASH have significantly larger total BA concentrations (Fig 10D) and conjugated-to-unconjugated BA ratios (Fig 10E) than cohorts with NAFL (p=0.0473, p=0.0192 respectively). In some studies, stages of NAFLD progression were not separated, and are presented here as NAFLD. Meanwhile, there are no differences in the secondary-to-primary BA ratios between cohorts with different stages of NAFLD progression (Fig 10F). There is insufficient data regarding cirrhotic cohorts, though they may have elevated total BA concentrations, but more evidence is needed. In all cases where there are significant differences, there is still an overlap between the two groups being compared, as well as with normal cohorts suggesting that successfully classifying a disease based on one of these metrics has a high probability of being inaccurate. Filtering the data for the recommendations to reduce variability (Table 3) reveals that data collected in accordance with the recommendations is limited to three datasets for normal cohorts, and three for metabolic liver diseases (NAFLD+Cirrhosis), with no datasets available for other diseases (Fig 11).

**Fig 11.**
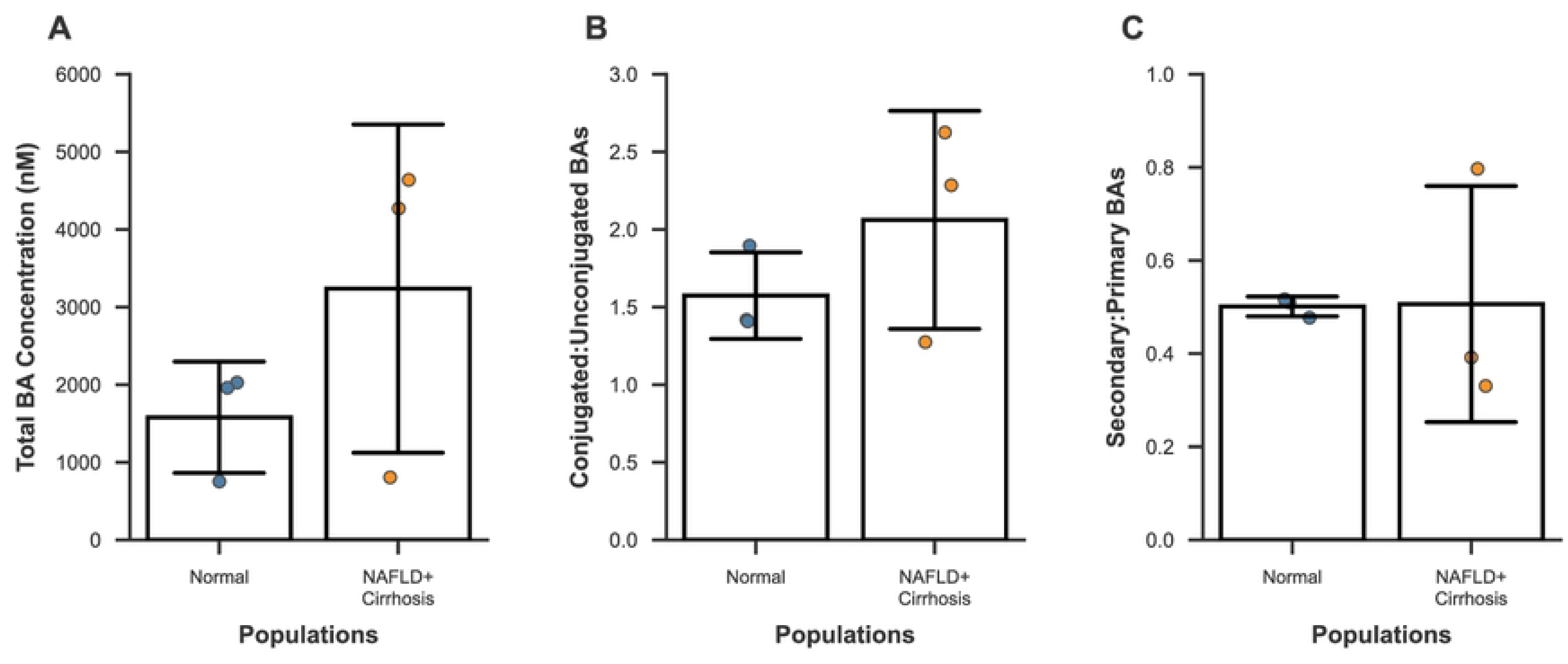
Data collected in accordance with the suggested recommendations are limited. The mean/median of the total bile acid concentrations (A), conjugated-to-unconjugated bile acid ratios (B) and secondary-to-primary bile acid ratios (C) in normal cohorts (those without enterohepatic disorders) and cohorts with metabolic-dysfunction associated liver disorders (NAFLD + cirrhosis) in which methanol was used as the protein precipitation solvent and LC-MS was used to measure bile acid concentrations in 50µl of plasma obtained from subjects fasted at the time of sampling were plotted as their mean ± standard deviation.

### Individual BA proportions in peripheral blood are significantly altered in enterohepatic disorders

When analysed using LC-MS, individual BAs can be reported as a proportion of the total BA concentration. While absolute BA concentrations are reported in all the included studies, the potential of BA proportions as a biomarker of health status was investigated. 47 datasets provide individual BA concentrations for the 15 BAs we focussed on in this study. 15 of these datasets are from normal cohorts, while the remaining 32 are from unhealthy cohorts with enterohepatic disorders. 14 of these 32 are from cohorts with metabolic-dysfunction associated liver diseases (NAFL, NASH & Cirrhosis), with the other 18 from cohorts with other liver diseases such as viral hepatitis, ALD, HCC (3 cohorts) and cholestasis (1 cohort). For each dataset, the 15 individual BA concentrations were summed for the total BA concentration, and the proportion of each individual BA of this total was determined. In these 47 cohorts, circulating concentrations of CA (p=0.0046), CDCA (p=0.0325), G-CDCA (p=0.0100), G-DCA (p=0.0060), G-LCA (p=0.0366), T-CDCA (p=0.0374) and T-LCA (p=0.0132) were significantly elevated in unhealthy cohorts compared to normal cohorts (Fig 12A). Statistically there is no significant difference in the concentrations of individual BAs between cohorts with other, non-metabolic dysfunction associated enterohepatic disorders and cohorts with metabolic-dysfunction associated enterohepatic disorders (S8 Fig A). Between normal cohorts and cohorts with metabolic-dysfunction associated enterohepatic disorders, individual BA concentrations remain similar (S9 Fig A).

**Fig 12.**
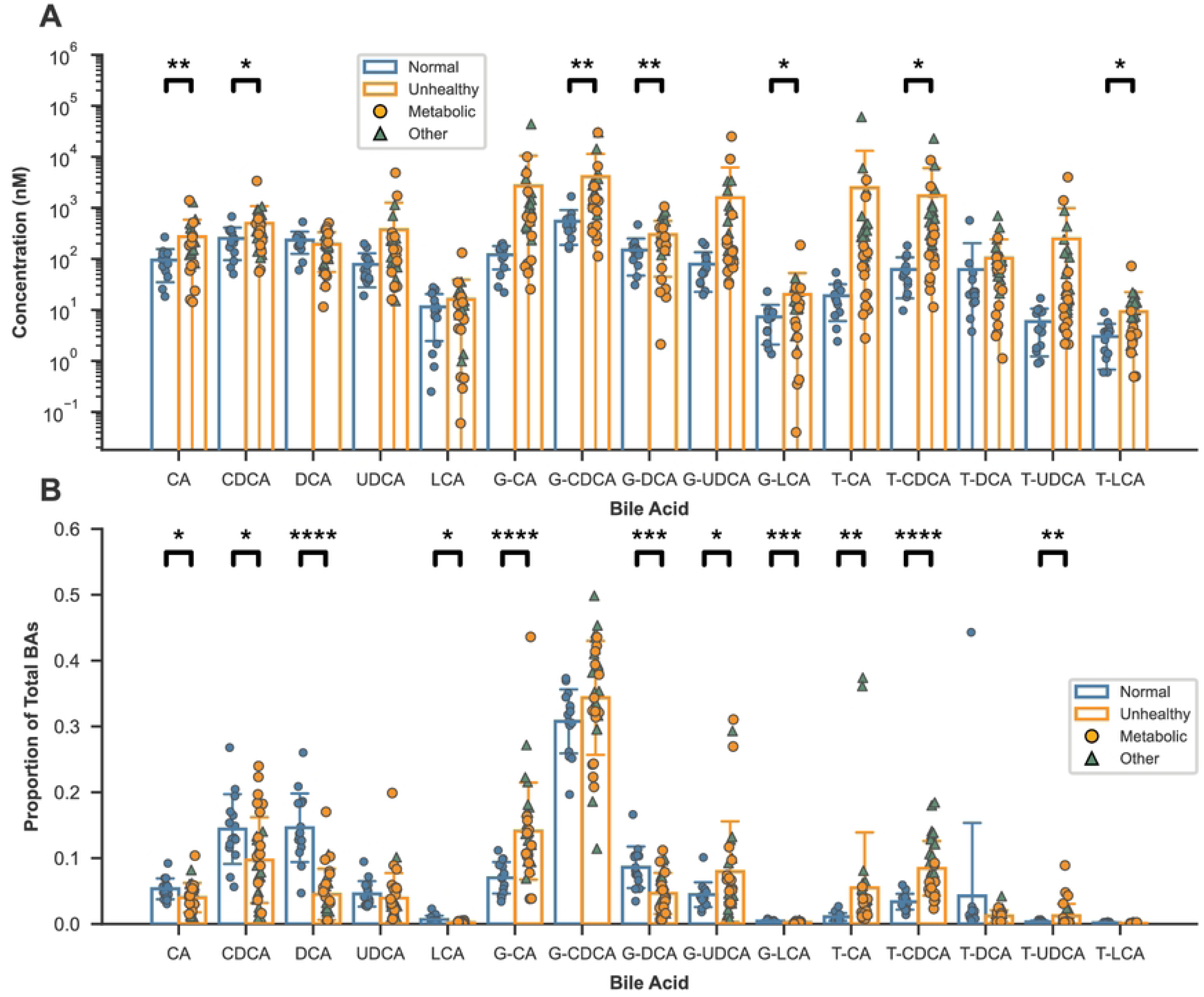
Circulating individual bile acids as a proportion of total bile acid concentrations are significantly altered in populations with enterohepatic disorders. The mean/median concentrations of peripheral bile acids from 47 datasets in which concentrations of the 15 bile acids focussed on in this study were reported in normal populations (without enterohepatic disorders) and unhealthy populations (with enterohepatic disorders) were plotted as their mean ± standard deviation (A). For each dataset, individual bile acid concentrations were totalled, and individual bile acids were plotted as their proportion of the total (B). Unhealthy populations were split into two groups: (1) Metabolic – populations with metabolic-dysfunction associated liver disorders (non-alcoholic fatty liver, non-alcoholic steatohepatitis, cirrhosis), and (2) Other – populations with other enterohepatic disorders. Statistics performed by two-tailed unpaired Welch’s t-test, blank p>0.05, * p≤0.05, ** p≤0.01, *** p≤0.001, **** p≤0.0001.

In contrast, the proportions of BAs are significantly altered. The proportions of CA (p=0.0234), CDCA (p=0.0126), DCA (p=0.000001), LCA (p=0.0101), G-DCA (p=0.0004), and G-LCA (p=0.0005) of total BAs are decreased in unhealthy cohorts (Fig 12B). Of these, the proportions of CDCA (p=0.0074), DCA (p=0.0239) and UDCA (p=0.0195) are significantly lower in cohorts with other enterohepatic disorders compared to metabolic-dysfunction associated enterohepatic disorders (S8 Fig B). Proportions of DCA (p=0.000075), LCA (p=0.0128), G-DCA (p=0.0013) and G-LCA (p=0.0003) are significantly decreased in normal cohorts compared to cohorts with metabolic-dysfunction associated disorders (S9 Fig B). Meanwhile, the proportions of G-CA (p=0.00001), G-UDCA (p=0.0196), T-CA (p=0.0061), T-CDCA (p=0.0000001) and T-UDCA (p=0.0093) of total BAs are increased in unhealthy cohorts. Of these, T-CA (p=0.0339) and T-CDCA (p=0.00003) proportions are significantly elevated in cohorts with metabolic-dysfunction associated enterohepatic disorders compared to other disorders and are significantly higher in cohorts with metabolic-dysfunction associated enterohepatic disorders than in normal cohorts (T-CA: p=0.0121; T-CDCA=0.0023).

## Discussion

This meta-analysis aimed to assess the variability range in peripheral BA concentrations reported in literature and to identify the biological, experimental and analytical determinants influencing this variability. A total of 65 studies, encompassing 10,629 individuals, were included in the analysis.

As expected, LC-MS was found to be the predominantly used analytical technique to measure individual BA concentrations. A comparison of individual BA concentrations measured using LC-MS and those measured using GC-MS in normal cohorts was not conclusive due to limited GC-MS data (S2 Fig). However, a comparison of totalBAs by both techniques revealed that concentrations of totalCA, totalCDCA and totalDCA measured using GC-MS are higher on average. These differences could be a consequence of calculating the LC-MS total BA concentrations by summing the concentrations of the unconjugated form, and the G- and T-conjugated forms (e.g. totalCA = CA + G-CA + T-CA), ignoring other forms such as sulphated BAs. Meanwhile, analysis by GC-MS requires the cleavage of all conjugates (79). The concentrations of the resulting unconjugated BAs are then measured. Hence, total BA concentrations measured using GC-MS are expected to be more accurate and higher than those measured using LC-MS, because all forms of BAs will be included, including sulphated forms.

On average, BA concentrations are found to be higher in cohorts fed at the time of sampling. Postprandial stimulation of BA secretion from the gallbladder into the small intestine to aid in the digestion and absorption of dietary fats results in an increase in BA pool size in the EHC, and consequently an increase in circulating BA concentrations due to leakage into the system circulation. In order to accurately assess inter-individual variability, it would be beneficial to compare samples obtained in the fasted state (ideally 12 hours) since preoperative patients follow *nil per os* guidelines. If the response to feeding is of interest, blood collection should be performed at a defined and consistent time following the ingestion of a standardized meal in otherwise fasting individuals. These conditions should be experimentally validated to account for variability due to differences in gallbladder filling and emptying.

Another source of variability in BA concentrations is the choice of blood collection matrix: serum or plasma. Serum samples tend to yield higher BA concentrations than plasma samples, while variability seems lower in plasma samples. Logistically, plasma is easier and faster to prepare, since no clotting time is required. Plasma also has a longer shelf-life compared to serum (years compared to months) (80). Studies have shown that concentrations of (non-BA) metabolites in serum and plasma drawn from the same patients can be significantly different (81,82). One of these study found the metabolite concentrations measured to be higher in serum than in plasma (82). The authors speculate that this observation could partly be a consequence of the volume displacement effect, where the deproteinization of serum samples eliminates a larger volume of proteins, leaving the metabolites in a smaller volume, making them more concentrated (83). However, plasma should contain similar, if not larger volumes of proteins than serum, due to the presence of coagulation factors such as fibrinogens (84). Therefore, the volume left after deproteinization should in fact be smaller in plasma samples, and so plasma should have higher metabolite concentrations. It should be noted that concentration differences between the two matrices were not consistent, with some metabolites having higher concentrations in serum and others having higher concentrations in plasma. Although both these studies used analytical techniques that were not LC-MS (81,82) (^1^H-NMR (Nuclear Magnetic Resonance Spectroscopy and Flow Injection Analysis-Mass Spectrometry), and BAs were not included in the metabolites quantified, it is reasonable to assume that there could be a difference in LC-MS BA concentrations measured in serum and in plasma obtained from the same patient. A study comparing the quantification of antidepressants by LC-MS in whole blood, plasma and serum found that using the calibration curve generated in one of the three matrices to quantify an analyte in the other two matrices was inconsistent and only partially successful (85), illustrating that the blood collection matrix is a source of variability, and should therefore be standardized to aid valid clinical interpretation. Additionally, the choice of anticoagulant has an impact on metabolite concentrations measured in plasma (81). Further investigation is required to better understand the use of serum or plasma from the same patient affects BA concentrations measured using LC-MS and the underlying reasons for concentration discrepancies between the two matrices. The relative logistical ease of obtaining plasma, and the higher reproducibility of metabolite concentrations in plasma (81,82) suggest that plasma is the better choice of matrix.

Plasma samples must be processed before analysis by LC-MS. Since the target BAs are present at low levels in relation to other plasma components, they must be selectively isolated and concentrated to increase the sensitivity of LC-MS. It is a possibility that when too low volumes of plasma are concentrated, the remaining amounts of low abundant BAs may be below the limit of detection of LC-MS instruments and may be falsely considered absent from the BA pool. Hence, the starting volume of plasma to be analysed is important to take into consideration to improve the comparability of studies. 50µl of plasma is sufficient to detect a wide range of concentrations for most BAs and is comparable to 100µl of plasma. BA concentration data collected from volumes of plasma less than 50µl is limited and is an area that could be investigated further. Thus, we recommend using 50µl of plasma to ensure accurate absolute BA concentration measurements whilst keeping a reasonably low volume of blood from a subject.

Another key aspect of sample preparation is the removal of highly abundant proteins by protein precipitation. The choice of solvent used for protein precipitation can influence the variability of measured BA concentrations. Methanol appears to be the best solvent since mean T-BA concentrations in plasma samples are higher than mean T-BA concentrations when acetonitrile was used. However, comparable data is limited, making it difficult to conclude which solvent is truly better to use to reduce variability. A comparison of extraction techniques followed by HPLC-MS/MS found that acetonitrile precipitation followed by C_18_-Solid Phase Extraction (SPE) was the most suitable technique for serum extraction of cBAs and uBAs (45). Additionally, methanol precipitation without SPE was able to detect the same 17 BAs but to a lower level. SPE uses a cartridge-bound stationary phase sorbent to selectively retain either target analytes or interfering substances and has high selectivity, high analyte recovery and high ease of automation (79,86,87). BA concentrations where SPE was used are scarce. The use of SPE in the LC-MS analysis pipeline could be crucial to maximising the accuracy of BA concentration measurements and warrants further investigation.

To reduce variability and improve comparability between studies, it is essential to standardise methodologies, including fasting status, sample type, analytical technique, sample volume, and protein precipitation methods. There is a lack of studies utilising the proposed standardised methodologies making a consideration of disease states difficult. Without consideration for whether studies used the suggested standardised methodologies, peripheral BA proportions appear to be significantly altered in subjects with an enterohepatic disorder. While standardised methodologies are important for ensuring accuracy and reproducibility, peripheral BA proportions, even when measured using various methods, could still provide valuable information for the diagnosis and monitoring of enterohepatic diseases. While circulating BAs hold potential as biomarkers of enterohepatic disorders, access to the BA concentration or proportions in individual subjects rather than cohort means, is critical for advancing this line of research. Additionally, standardized sampling, sample preparation, and analysis are prerequisites for future studies aiming to uncover the diagnosis or prognosis value of circulating BAs. This would help to understand how variable inter-individuals circulating BA concentrations or proportions are, which would be particularly useful in situations where the availability of data for some diseases, such as cirrhosis, is limited, hindering a comprehensive analysis of the diagnostic and prognostic value of circulating BAs.

The main limitation of this study is that the determinants of variability in BA concentrations were identified both across studies and across cohorts. To confirm whether these are true sources of variability and not some artefact of biased data, studies investigating those determinants in samples collected from the same patients would be needed.

In conclusion, this meta-analysis provides a comprehensive overview of the biological, experimental and analytical determinants influencing BA concentration variability in human peripheral blood and highlights the importance of standardization in future research. By addressing these limitations and conducting further studies, we can hope to rely on BA as relevant biomarkers for liver diseases, especially MASLD severity.

## Data Availability

All bile acids data files are available from the github database (https://github.com/sebjoseph122/Bile-Acid-Literature-Review).

## Supporting information

**S1 Fig.** Mean/Median total bile acid concentrations (A), the concentrations of the most abundant bile acid: G-CDCA (B) and the concentrations of the least abundant bile acid: T-UDCA (C) in normal cohorts (those without enterohepatic disorders) were plotted against the average age of the respective cohort. The blue lines are the regression estimate and the shaded blue regions are the 95% confidence interval for the regression estimate.

**S2 Fig.** Mean/Median individual bile acid concentrations (A) and total bile acid concentrations (B) in normal cohorts (those without enterohepatic disorders) were stratified for whether LC-MS or GC-MS were used to measure the bile acid concentrations and plotted as their mean ± standard deviation. Statistics performed by two-tailed unpaired Welch’s t-test, blank p>0.05, * p≤0.05, ** p≤0.01, *** p≤0.001.

**S3 Table.** A comparison of the variance of serum and plasma peripheral bile acid concentrations in normal cohorts fasted at the time of sampling from studies using LC-MS as the analytical technique.

**S4 Fig.** Mean/median concentrations of the most abundant bile acid: G-CDCA (A) and the least abundant bile acid: T-UDCA (B) from studies that used LC-MS in plasma obtained from normal cohorts (those without enterohepatic disorders) which were fasted at the time of sampling were plotted against the volume of plasma used for analysis.

**S5 Fig.** The protein precipitation solvent used in datasets from studies that used LC-MS to measure bile acid concentrations in 50µL and 100µL plasma from normal cohorts (those without enterohepatic disorders) which were fasted at the time of sampling were tallied and presented as a pie chart (A). The total bile acid concentrations of these datasets were summed and stratified for the protein precipitation solvent used and plotted as their mean ± standard deviation (B). The mean/median concentrations of glycine-conjugate bile acids (C) and taurine-conjugate (D) were stratified for the protein precipitation solvent used and plotted as their mean ± standard deviation.

**S6 Fig.** Mean/Median individual bile acid concentrations (A) and total bile acid concentrations (B) in normal cohorts (those without enterohepatic disorders) were collected from literature and compared to those in filtered normal cohorts i.e. those from studies in which bile acid concentrations in 50µl of plasma obtained from patients fasted at the time of sampling were measured using LC-MS, and methanol was used as the protein precipitation solvent. Both groups were plotted as their mean ± standard deviation.

**S7 Table.** A comparison of the variance of peripheral bile acid concentrations in all normal cohorts and filter normal cohorts i.e. those from studies in which bile acid concentrations in 50µl of plasma obtained from patients fasted at the time of sampling were measured using LC-MS, and methanol was used as the protein precipitation solvent.

**S8 Fig.** The mean/median concentrations of peripheral bile acids from 32 datasets in which concentrations of the 15 bile acids focussed on in this study were reported in cohorts with metabolic-dysfunction associated liver disorders (non-alcoholic fatty liver, non-alcoholic steatohepatitis, cirrhosis), and cohorts with other enterohepatic disorders were plotted as their mean ± standard deviation (A). For each dataset, individual bile acid concentrations were totalled, and individual bile acids were plotted as their proportion of the total (B). Statistics performed by two-tailed unpaired Welch’s t-test, blank p>0.05, * p≤0.05, ** p≤0.01, **** p≤0.0001.

**S9 Fig.** The mean/median concentrations of peripheral bile acids from 29 datasets in which concentrations of the 15 bile acids focussed on in this study were reported in normal cohorts (those without enterohepatic disorders) and those with metabolic-dysfunction associated liver disorders (non-alcoholic fatty liver, non-alcoholic steatohepatitis, cirrhosis) were plotted as their mean ± standard deviation (A). For each dataset, individual bile acid concentrations were totalled, and individual bile acids were plotted as their proportion of the total (B). Statistics performed by two-tailed unpaired Welch’s t-test, blank p>0.05, * p≤0.05, ** p≤0.01, *** p≤0.001, **** p≤0.0001.

## Notes

### Competing Interest Statement

The authors have declared no competing interest.

### Funding Statement

Yes

